# Assessment of Early Mitigation Measures Against COVID-19 in Puerto Rico: March 15-May 15, 2020

**DOI:** 10.1101/2020.06.11.20127019

**Authors:** Miguel Valencia, José E. Becerra, Juan C. Reyes, Kenneth G. Castro

**Affiliations:** Data Management Operations for COVID-19 Legacy Systems, Puerto Rico Department of Health, San Juan, PR; Graduate School of Public Health, University of Puerto Rico Medical Sciences Campus, San Juan, PR; Department of Biostatistics and Epidemiology, Graduate School of Public Health, University of Puerto Rico Medical Sciences Campus, San Juan, PR; Hubert Department of Global Health & Department of Epidemiology, Rollins School of Public Health, Division of Infectious Diseases, Department of Medicine, School of Medicine, Emory University, Atlanta, GA

## Abstract

**Background:** On March 15, 2020 Puerto Rico implemented non-pharmaceutical interventions (NPIs), including a mandatory curfew, as part of a state of emergency declaration to mitigate the community transmission of the SARS-CoV-2 virus. The strict enforcement of this curfew was extended through May 25, with a gradual relaxation beginning on May 1. This report summarizes an assessment of these early mitigation measures on the progression of COVID-19 in the island.

**Methods and Findings:** From March 15 to May 15, 2020, 41,748 results of molecular (RT-PCR) tests were reported to the Puerto Rico Department of Health. Of these, 1,866 (4.5%) were positive, corresponding to 1,219 individuals with COVID-19 included in the study. We derived the epidemic growth rates (r) and the corresponding reproductive numbers (R) from the epidemic curve of these 1,219 individuals with laboratory-confirmed diagnosis of COVID-19 using their date of test collection as a proxy for symptoms onset. We then compared the observed cases with the R-based epidemic model projections had the mitigation measures not been implemented. Computations were conducted in the R packages forecast, incidence and projections.

The number of daily RT-PCR-confirmed cases peaked on March 30 (84 cases), showing a weekly cyclical trend, with lower counts on weekends and a decreasing secular trend since March 30. The initial exponential growth rate (r) was 17.0% (95% CI: 8.4%, 25.6%), corresponding to a doubling of cases every 4.1 days, and to a reproduction number (Ro) of 1.89 (95% CI: 1.41, 2.39). After March 30, the r value reverted to an exponential decay rate (negative) of −3.6% (95% CI: −5.7%, −1.4%), corresponding to a halving of cases every 19.4 days and to an Ro of 0.90 (95% CI: 0.84, 0.97). Had the initial growth rate been maintained, a total of 18,699 (96%CI: 4,113, 87,438) COVID-19 cases would have occurred by April 30 compared with 1,119 observed.

**Conclusions:** Our findings are consistent with very effective implementation of early non-pharmaceutical interventions (NPIs) as mitigation measures in Puerto Rico. These results serve as a baseline to assess the impact of the transition from mitigation to containment stages in Puerto Rico.

## Introduction

A novel coronavirus designated SARS-CoV-2 has been associated with coronavirus disease 2019 (COVID-19). The disease spectrum ranges from asymptomatic infections, mild-to-moderate influenza-like illness, pneumonia, severe acute respiratory distress, hyperinflammatory state, coagulopathy, to death.^1,2,3^ First described in late 2019 in China, this novel virus rapidly spread for person-to-person and cases were identified in other provinces and 19 additional countries. By January 30, 2020, the World Health Organization (WHO) declared the COVID-19 outbreak a global health emergency.^4^ With the rapid spread of this condition to a total of 113 countries, WHO Director General declared a global pandemic on 11 March 2020.^5^

In the absence of safe and effective vaccine or other therapeutic modalities against SARS-CoV-2, early measures in China included the implementation of non-pharmaceutical interventions (NPIs) aimed at temporizing the spread, illness, and deaths due to COVIV-19.^6^ Similar interventions have been implemented in other countries; NPIs had previously been successfully used to mitigate the 1918-1919 influenza pandemic.^7^ In the United States and Territories, the President and Governors declared health emergencies to implement NPIs, including orders to stay at home, travel restrictions, and closure of nonessential businesses.^8^

On March 13, 2020 the first confirmed COVID-19 cases were identified in Puerto Rico.^9^ These were two tourists from Italy (ages 68 and 70), and a 71-year-old Puerto Rican with lymphoma and diabetes, whose relatives had recently traveled to Chicago, Illinois. Within the past 30 months Puerto Rico had experienced several major natural disasters, including hurricanes Irma and Maria in September 2017 with widespread interruption of the infrastructure for essential services. By late December 2019 and January 2020, the island experienced significant earth tremors and multiple aftershocks with extensive structural damage and housing instability in the Southwest, associated with limited health care access among vulnerable populations.^10,11,12^ Against this backdrop, on March 15, 2020, the Governor of Puerto Rico issued an island-wide mandatory curfew as part of a state of emergency declaration closing public and private schools and universities, all nonessential businesses and public agencies. By then, five COVID-19 cases had been confirmed. The curfew, and its strict enforcement, were extended through May 25, with a gradual relaxation beginning on May 1.

## Methods

Using FDA-approved laboratory tests under Emergency Use Authority (EUA), all molecular tests by real-time polymerase chain reactions (RT-PCR) were conducted in approximately 900 laboratories throughout the island from March 15 through May 15, 2020 and reported to the Puerto Rico Department of Health. All positive RT-PCR test results were de-duplicated to account for individuals who had more than one clinical specimen submitted for testing, and to accurately derive the number of individual COVID-19 cases over time (by date of sample collection). Using the epidemic curve, we compared the pre- and post-estimates of the epidemic growth rate (r) and its corresponding reproductive number (Ro) by March 30, two weeks after the mandatory curfew was implemented. Deaths were included for analysis through May 22 to account for the censoring effect in the registration of recent deaths. Results of a comprehensive retrospective cohort study of close contacts in Shenzhen, China, were used to impute the serial interval distribution (mean=6.3 days, SD=4.2).^13^ Computations were conducted in the R packages forecast, incidence and projections.^14^

## Results

Among residents of Puerto Rico, a total of 46,093 results of molecular (RT-PCR) tests were reported to the Puerto Rico Department of Health between March 15 and May 15. Of these, 1,991 (4.3%) were positive, corresponding to 1,256 individuals with COVID-19. Demographically, 50.5% were male with a mean age of 45.9, and 49.5% female with a mean age of 45.5.

The number of daily RT-PCR confirmed cases peaked on March 30 (84 cases), showing a weekly cyclical trend, with lower counts on weekends and a decreasing secular trend since March 30 (Figure 1). The initial exponential growth rate (r) was 17.0% (95% CI: 8.4%, 25.6%), corresponding to a doubling of cases every 4.1 days, and to a reproduction number (Ro) of 1.89 (95% CI: 1.41, 2.39).

**Figure 1.**
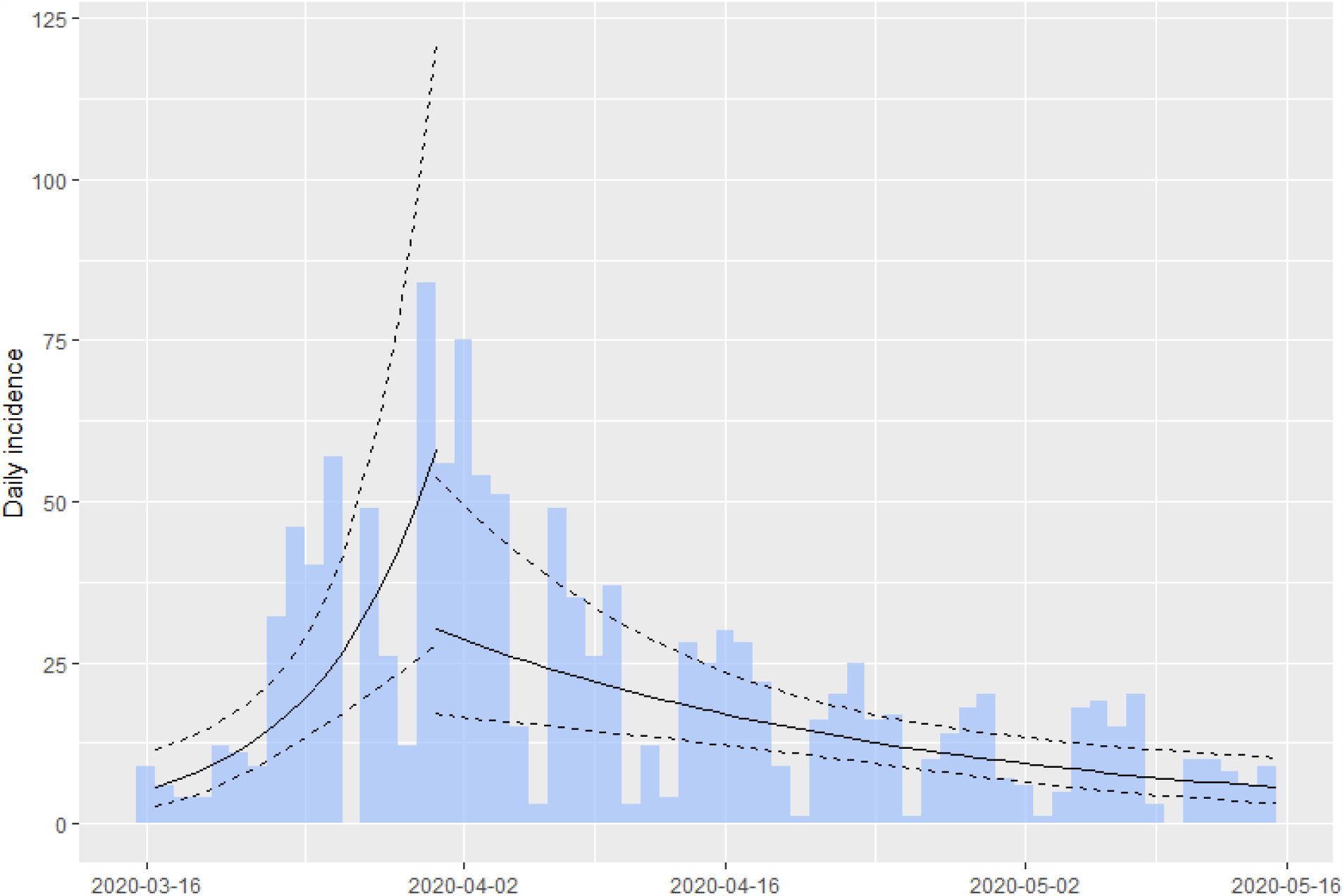
Daily incidence of COVID-19 cases (histograms), March 15–May 15, and epidemic curve showing pre and post March 30, 2020 exponential growth fits (with 95% CI)

After March 30, the r value reverted to an exponential decay rate (negative) of −3.6% (95% CI: −5.7%, −1.4%), corresponding to a halving of cases every 19.4 days and to an Ro of 0.90 (95% CI: 0.84, 0.97). Had the initial exponential growth rate been maintained, a total of 18,699 (96%CI: 4,113, 87,438) COVID-19 cases would have occurred by April 30 compared with 1,119 observed (Figure 2). A more conservative forecast using the linear growth rate from a seasonally adjusted Auto Regressive Integrated Moving Average (ARIMA) model (0,1,0) shows that 6,294 (95%CI: 5,575; 7,013) cases would have occurred in the same period.

**Figure 2.**
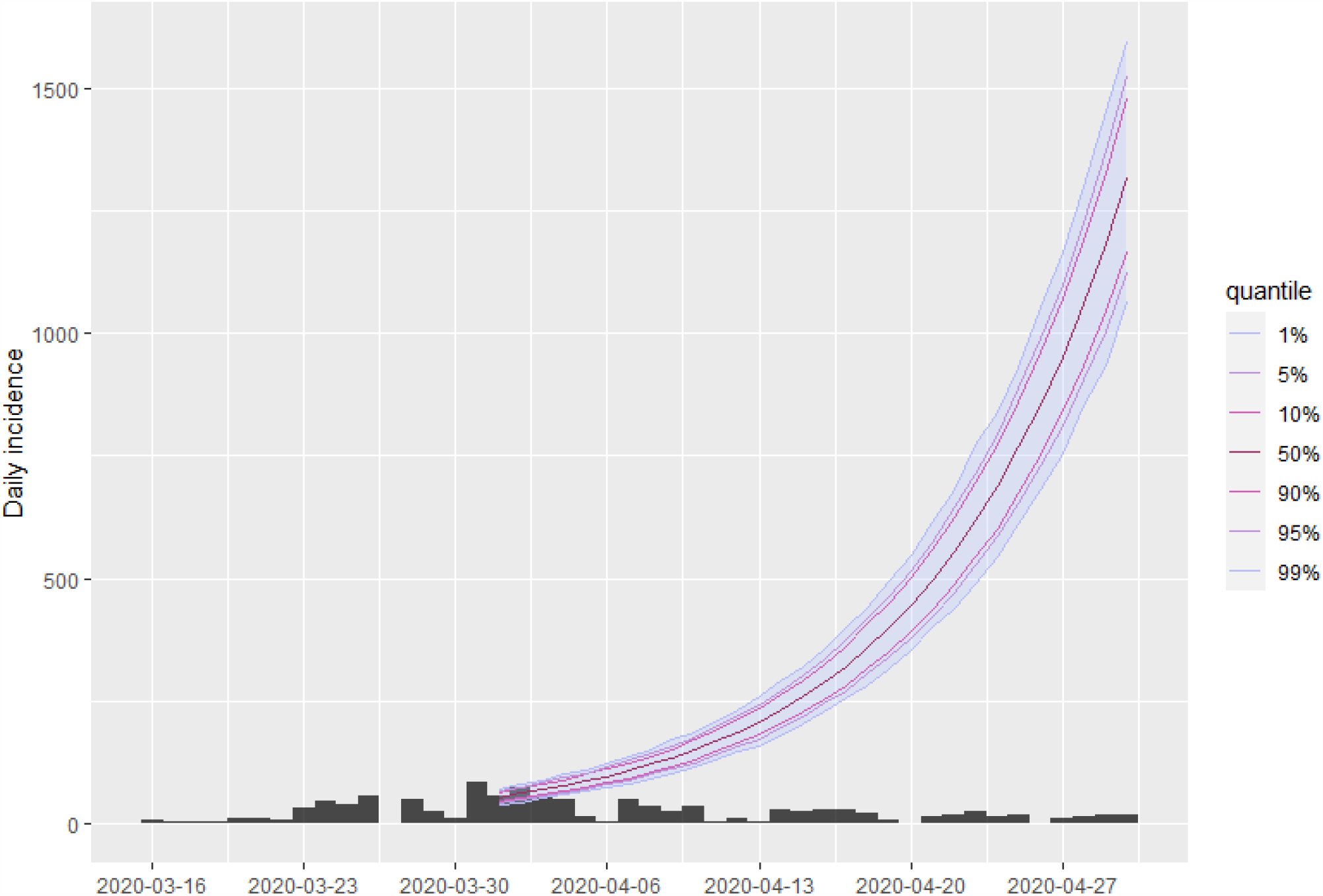
Projected daily incidence of COVID-19 cases through April 30, 2020

Throughout the observation period, the total number of RT-PCR tests per day fluctuated but trended to a lower proportion of positive results (from 16.2% in the first two weeks to 1.7% in the last two weeks of the study period).

A total of 132 COVID-19 deaths were reported from March 15 through May 22, 2020. Of these, 115 (87.1%) had RT-PCT test performed, 55 (41.7%) were laboratory confirmed and 77 (58.3%) had COVID-19 as a probable cause of death; 56.7% were male with a median age of 70.5, and 43.3% were female with a median age of 78.0.

The daily COVID-19 deaths peaked on April 13, 2020 (6 deaths). As illustrated in Figure 3, the initial exponential growth rate (r) of COVID-19 deaths was 5.9% (95% CI: 3.2%, 8.6%), corresponding to a doubling of deaths every 11.8 days. After April 13, the r value reverted to an exponential decay rate (negative) of −2.7% (95%CI: −4.3%, −1.3%), corresponding to a halving of cases every 25.0 days. An estimated additional 510 (95%CI: 130, 890) COVID-19 deaths would had occurred by May 22 if the initial exponential growth of 5.9% had not been curbed.

**Figure 3.**
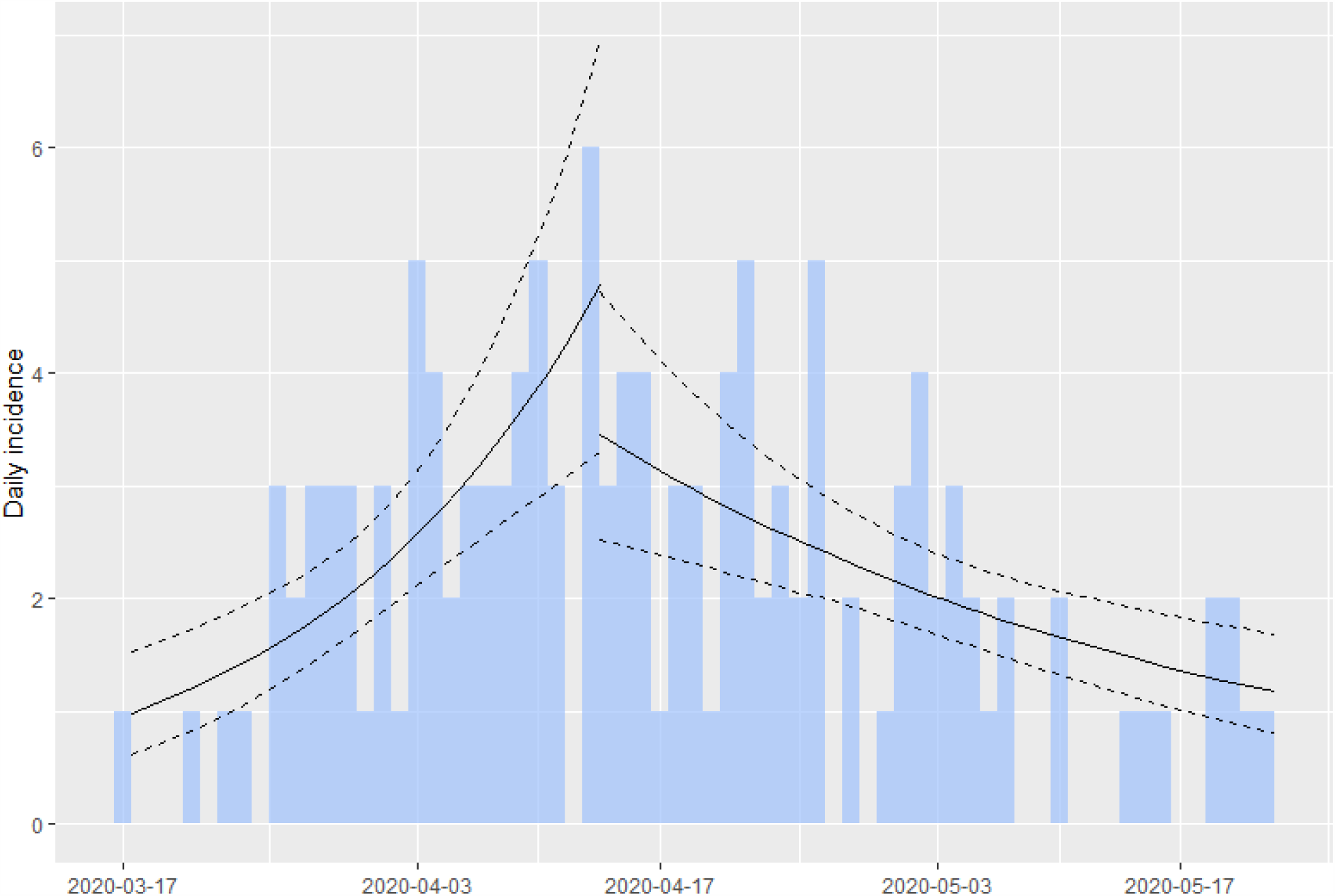
Daily COVID-19 deaths, March 15–May 22, and epidemic curve of 127 COVID-19 deaths from March 17 to May 22, 2020

## Discussion

Our findings are consistent with the use of NPIs as very effective early COVID-19 mitigation measures in Puerto Rico. It is likely that detection of COVID-19 cases remains under ascertained on the island, as suggested by the relatively high crude estimate of the case fatality rate of about 10%. However, this under ascertainment should not alter the estimates obtained from the epidemic curve provided the case ascertainment remained relatively constant throughout the study period. The decreasing trend in the proportion of RT-PCR positive tests lend support to this assumption.

The mortality curve is consistent with a 2-week delay relative to the epidemic curve of the COVID-19 cases, as well as with hospital data reported elsewhere,^9^ confirming our findings about the effectiveness of the mitigation efforts. COVID-19 mortality may also be under ascertained in Puerto Rico.

These results show benefits associated with the early implementation of NPIs for COVID-19, with stay-at home orders, accompanied by mandated curfew and closure of non-essential services in a manner consistent with historical reports of benefits observed with similar use of NPIS in various U.S. cities during the 1918-1919 influenza pandemic^7^ For sustained benefit, robust ongoing surveillance system will help identify incidence changes to inform policy decisions. In addition, these data serve as a baseline to assess the impact of the transition from mitigation to containment stages in Puerto Rico. We recommend that RT-PCT testing capacity remain available for all symptomatic individuals in Puerto Rico, as well as for all essential workers, and that the public health infrastructure for case investigation and contact tracing, accompanied by targeted isolation and quarantine, continue to be strengthened and scaled up to safeguard the gains observed to date.

## Data Availability

The public data used for our study was anonymized prior to our team having access. The Puerto Rico Department of Health anonymized the data. The public data is available at the URL below.
The specific subset of the pubic data set used for our analysis (including the R code) will be made available upon acceptance of manuscript by PLOS Medicine.

https://estadisticas.pr/en/covid-19

## Notes

### Competing Interest Statement

The authors have declared no competing interest.

### Funding Statement

no external funding was received

### Author Declarations

Category of exemption: D. Research involving the collection or study of existing data, documents, records, pathological specimens, or diagnostic specimens, if these sources are publicly available or if the information is recorded by the investigator in such a manner that subjects cannot be identified, directly or through identifiers linked to the subjects. https://irbrcm.rcm.upr.edu/wp-content/uploads/sites/21/2020/04/Procedure-for-Determining-Exemptions.pdf

